# Can individuals with suboptimal antibody responses to conventional antiviral vaccines acquire adequate antibodies from SARS-CoV-2 mRNA vaccination?

**DOI:** 10.1101/2021.12.26.21268358

**Authors:** Wataru Ogura, Kouki Ohtsuka, Sachiko Matsuura, Takahiro Okuyama, Satsuki Matsushima, Satoko Ymasaki, Hiroyuki Miyagi, Kumiko Sekiguchi, Hiroaki Ohnishi, Takashi Watanabe

## Abstract

**Objective:** In Japan, healthcare workers (HCWs) are vaccinated against contagious viruses (measles, rubella, chickenpox, mumps, and hepatitis B) to prevent nosocomial infection; however, some do not produce sufficient antibodies (suboptimal responders). Whether suboptimal responders to live attenuated viruses or inactivated viruses vaccines can produce adequate antibodies to severe acute respiratory syndrome coronavirus 2 (SARS-CoV-2) mRNA vaccines remains to be elucidated.

**Methods:** In this prospective cohort study, SARS-CoV-2 anti-spike antibodies were measured 11 times, from before the first BNT162b2 vaccination to 5 months after the second vaccination. Antibody titers of suboptimal and normal responders were compared. SARS-CoV-2 neutralizing antibody activity was measured twice in suboptimal responders, 1 week to 1 month, and 5 months after the second vaccination.

**Patients:** This study included 50 HCWs who received two doses of mRNA BNT162b2 vaccine 3 weeks apart.

**Results:** After vaccination, the SARS-CoV-2 anti-spike antibody was detectable in the samples from suboptimal and normal responders at each timepoint. The median SARS-CoV-2 anti-spike antibody titer was higher in suboptimal responders than in normal responders 1 week after receiving the second dose of BNT162b2 vaccine (3721.0 vs. 2251.5, *P*=0.029). Suboptimal responders had SARS-CoV-2 neutralizing antibody activity 1 week to 1 month, and 5 months after the second vaccination, which exceeded the positive threshold 5 months after the second vaccination.

**Conclusion:** After BNT162b2 vaccination, suboptimal responders acquired adequate SARS-CoV-2 anti-spike and SARS-CoV-2 neutralizing antibodies to prevent SARS-CoV-2. These results suggest that vaccination with mRNA vaccine against SARS-CoV-2 should also be recommended for suboptimal responders to conventional vaccines.

## Introduction

Coronavirus disease (COVID-19), which is caused by severe acute respiratory syndrome coronavirus 2 (SARS-CoV-2), has been declared a pandemic (1-4). In view of the high number of cases of severe illness and death due to COVID-19, vaccination against SARS-CoV-2 infection is crucial.

SARS-CoV-2 has an enveloped, single, positive-stranded RNA genome that encodes four major viral structural proteins, namely, the nucleocapsid, spike, envelope, and membrane proteins; the latter three proteins are found in its membrane. The spike protein guides viral entry into host cells by binding to angiotensin-converting enzyme 2 (ACE2) receptors, which are widely expressed on epithelial cells and macrophages (5-7) and is thus an ideal target for messenger RNA (mRNA) vaccine development.

The BNT162b2 mRNA vaccine (Pfizer–BioNTech) is highly effective at preventing clinically significant COVID-19 (8-10), reducing the incidence of asymptomatic infection and associated infectivity (11,12), and preventing the spread of COVID-19 (9). The BNT162b2 vaccine was the first SARS-CoV-2 vaccine to be approved in Japan, and priority vaccination of healthcare workers (HCWs) began in February 2021 in order to safeguard the medical delivery system (13).

HCWs also have a high risk of infection with other contagious viruses such as measles, rubella, chickenpox, mumps, and hepatitis B. Therefore, in Japan, HCWs are vaccinated against these viruses to prevent nosocomial infections and viral antibody titers are measured to ensure the presence of protective antibody levels (14). However, some individuals do not develop adequate antibody titers against contagious viruses even after vaccination. Such individuals are called suboptimal responders. It is unclear whether suboptimal responders to conventional vaccines can produce adequate SARS-CoV-2 antibodies, including neutralizing antibodies, after vaccination with the BNT162b2 vaccine to protect them against SARS-CoV-2 infection. Therefore, in this study, we investigated whether suboptimal responders acquired SARS-CoV-2 anti-spike and SARS-CoV-2 neutralizing antibodies after vaccination with the BNT162b2 vaccine.

## Materials and methods

### Participants

The study was conducted at Kyorin University Hospital in Tokyo, Japan. HCWs at the hospital received two doses of the BNT162b2 vaccine 3 weeks apart. The participants were HCWs at the hospital who agreed to participate in the study and to periodically donate blood samples before and after vaccination. Those whose antibody levels were below the standard values for preventing infection after receiving two or more doses of vaccine against measles, rubella, chickenpox, mumps, or hepatitis B viruses were classified as suboptimal responders.

### SARS-CoV-2 anti-spike antibody assay

SARS-CoV-2 anti-spike antibody measurements were performed on serum obtained from centrifuged peripheral blood using the Elecsys Anti-SARS-CoV-2 S RUO (Roche Diagnostics International Ltd, Rotkreuz, Switzerland) on a Cobas 6000 (Roche Diagnostics) analyzer via the double-antigen sandwich enzyme-linked immunoassay method (15). Samples were collected before the first dose of BNT162b2 vaccine was administered; 3 days after the first vaccination; 1, 2, and 3 weeks after the first vaccination; 1 week after the second vaccination; and 1, 2, 3, 4, and 5 months after the second vaccination. The measurement range of the antibody assay was 0.4–250, and samples with an antibody titer of ≥250 U/mL were assayed again after dilution.

### SARS-CoV-2 neutralizing antibody assay

Using serum obtained from the peripheral blood of suboptimal responders after vaccination, SARS-CoV-2 neutralizing antibody activity was measured at Osaka University Microbial Disease Research Association Laboratory. A 50% inhibitory dilution factor (ID_50_) using pseudovirus (16) was measured, and ID_50_ values of ≥50 was considered positive (17). For the neutralization assay, 60 μL of pseudovirus, which is equivalent to 2.5×10^6^ relative light units (RLU)/mL, was incubated with 60 μL of serial dilutions of serum samples for 1 h at 37LJ. After incubation, 100 μL of the mixture was added to the Vero cells in a 96-well plate. The cells were lysed 24 h post-infection, luciferase was activated using the Luciferase Assay System (E1501, Promega, Wisconsin, USA), and RLU were measured using a Synergy LX analyzer (BioTek, USA). Percent neutralization was calculated using GraphPad Prism 8.0.2 (GraphPad Software, San Diego, California, USA). Percentages of RLU reduction (inhibition rate) were calculated as follows: 1 − (RLU of sample sera – pseudovirus only wells) / (RLU from medium only wells − pseudovirus only wells) × 100%. The titers of neutralizing sera were calculated as ID_50_. The transition of neutralizing antibody activity values after vaccination in suboptimal responders was examined 1 week to 1 month and 5 months after receiving the second dose of the BNT162b2 vaccine.

### Statistical analysis

Nonparametric continuous data are reported as median and interquartile range (IQR). The Mann–Whitney U test was performed to assess whether differences between cohorts were statistically significant. Two-sided *P*-values of <0.05 were considered statistically significant. The Wilcoxon matched-pairs signed rank test was performed to assess whether differences in antibody levels within an individual at different time points were statistically significant. To compare the age (ratio of the number of individuals aged 20–40 years and those aged 50–60 years) and sex compositions of the suboptimal responders and normal responders, Fisher’s exact test was performed to test the statistical significance. The Mann–Whitney U test was performed to test for statistically significant differences in antibody levels between suboptimal responders and normal responders 1 week after the second vaccination. All statistical analyses were performed with EZR (Saitama Medical Center, Jichi Medical University, Saitama, Japan), which is a graphical user interface for R (The R Foundation for Statistical Computing, Vienna, Austria) (18). Specifically, EZR is a modified version of R commander (version 1.54) that has been designed to add statistical functions frequently used in biostatistics.

## Results

The suboptimal responders comprised three men (L1, L2, L4) with negative (titer <16.0) anti-measles immunoglobulin G (IgG) antibody titer after measles virus vaccination and one woman (L3) with a negative (titer <4.0) anti-mumps IgG antibody titer after mumps virus vaccination. Forty-six participants, comprising 12 men and 34 women, had antiviral antibody titers after vaccination against other viruses above the standard value (normal responders). The four suboptimal responders were aged 20–40 years. Normal responders were older than suboptimal responders (the ratios of the number of those aged 50–60 years were 57% vs. 0%, respectively), but no significant difference was observed (*P* >0.99). In addition, the male/female ratio was higher in suboptimal responders than in normal responders (74% vs. 15%), but no significant difference was observed (*P* >0.99). All suboptimal responders were non-smokers and non-obese, and none of them had a history of immunosuppressive drug use or having a medical condition associated with immunodeficiency.

### SARS-CoV-2 anti-spike antibody levels

The median (IQR) SARS-CoV-2 anti-spike antibody titer was 2251.5 (1639.0– 3276.3) in the normal responders and 3721.0 (3255.0–4983.3) in the suboptimal responders 1 week after receiving the second dose of BNT162b2 vaccine. This difference was statistically significant (*P*=0.029) (Fig. 1A). For the total antibody level 1 week after the second vaccination, there was no significant difference in the median antibody levels between those aged 50–60 years and those aged 20–40 years(*P*=0.66).

**Fig 1.**
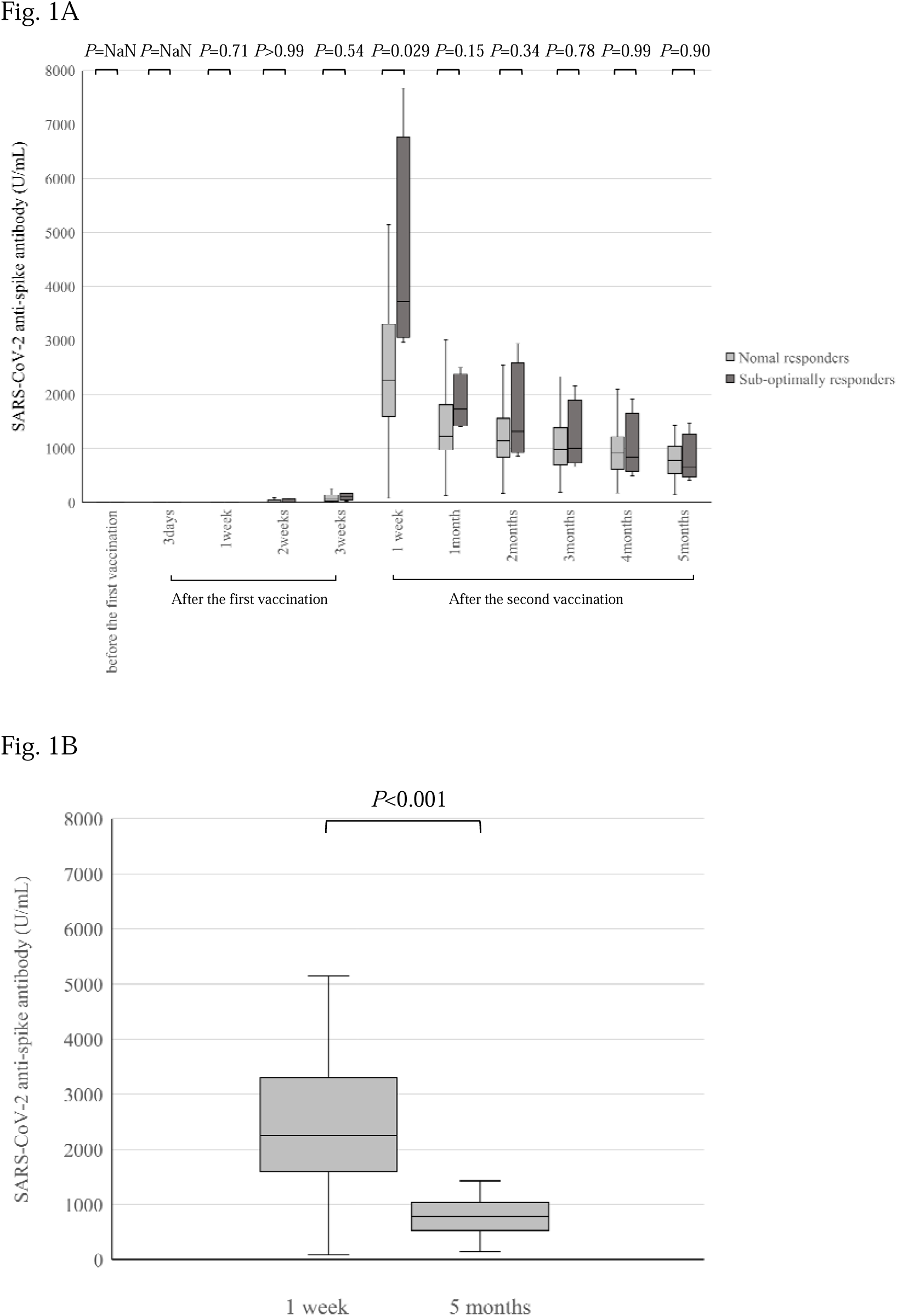
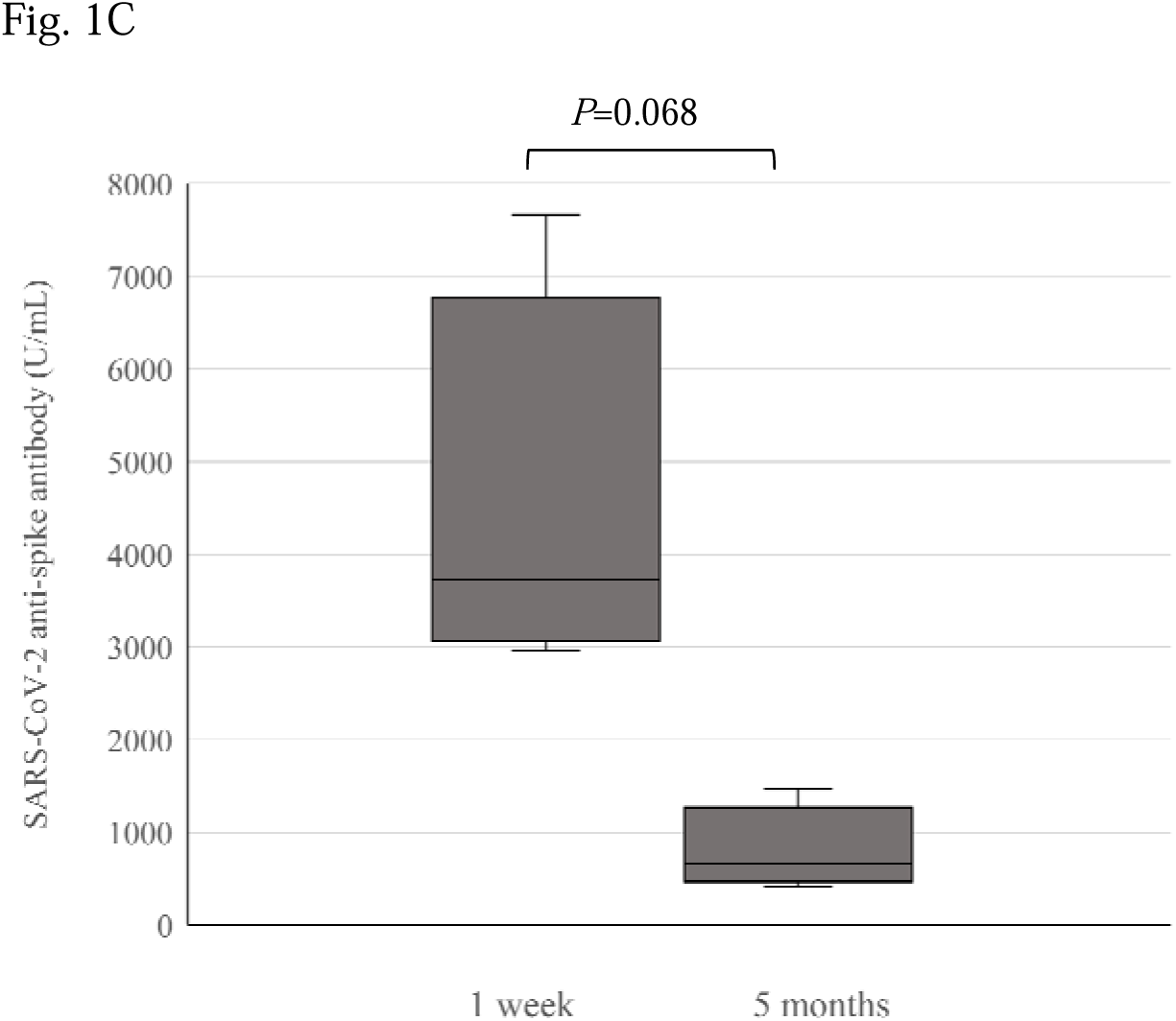
SARS-CoV-2 anti-spike antibody responses in suboptimal/normal responders before and after vaccination with the BNT162b2 vaccine.

The median (IQR) SARS-CoV-2 anti-spike antibody in normal responders was 2251.5 (1639.0–3276.3) and 779.3 (534.6–1024.3) measured 1 week and 5 months, respectively, after receiving the second dose of BNT162b2 vaccine (Fig. 1B). In suboptimal responders, the median (IQR) SARS-CoV-2 anti-spike antibody titters were 3721.0 (3255.0–4983.3) and 666.7 (578.9–880.6) measured 1 week and 5 months, respectively, after receiving the second dose of BNT162b2 vaccine (Fig. 1C).

SARS-CoV-2 anti-spike antibody was produced by both suboptimal and normal responders, but the level was lower at 5 months than at 1 week after the second dose of vaccine (Fig. 1).

A. SARS-CoV-2 anti-spike antibody responses over the whole study period from before the first dose to 5 months after the second dose of vaccine.
B. SARS-CoV-2 anti-spike antibody titers in normal responders 1 week and 5 months after receiving the second dose of the BNT162b2 vaccine. The antibody titers were significantly lower at 5 months than at 1 week after the second dose (*P*<0.001).
C. SARS-CoV-2 anti-spike antibody titers in suboptimal responders 1 week and 5 months after receiving the second dose of the BNT162b2 vaccine. The antibody titers decreased, but the decrease was not statistically significant (*P*=0.068).

The box plot comparisons were performed using the Mann–Whitney U test. *P*-values <0.05 were considered statistically significant. Suboptimal responders had low-level antibody responses to previous measles or mumps vaccines, and normal responders had normal antibody responses to previous antiviral vaccines.

NaN; not a number

### SARS-CoV-2 neutralizing antibody activity in participants with suboptimal antibody responses to previous antiviral vaccines

SARS-CoV-2 neutralizing antibody titers were 1154.0 in L1 at 1 week after receiving the second dose of BNT162b2 vaccine and 774.2, 638.3, and 654.6 in L2, L3, and L4, respectively, at 1 month after receiving the second dose of BNT162b2 vaccine (Fig. 2). In the 5 months after the second vaccination, SARS-CoV-2 neutralizing antibody titers were 105.5, 147.7, 485.2, and 894.4 in L1, L2, L3, and L4, respectively (Fig. 2). All four suboptimal responders had a neutralizing antibody activity within the protective range (≥50), from the 1 week to 1 month and 5 months after the second vaccination (Fig. 2). The SARS-CoV-2 neutralizing antibody level in suboptimal responders was lower at 5 months after receiving the second dose of BNT162b2 vaccine than at 1 week to 1 month after receiving the second dose, but this difference was not statistically significant (*P*=0.38) (Fig. 2).

**Fig 2.**
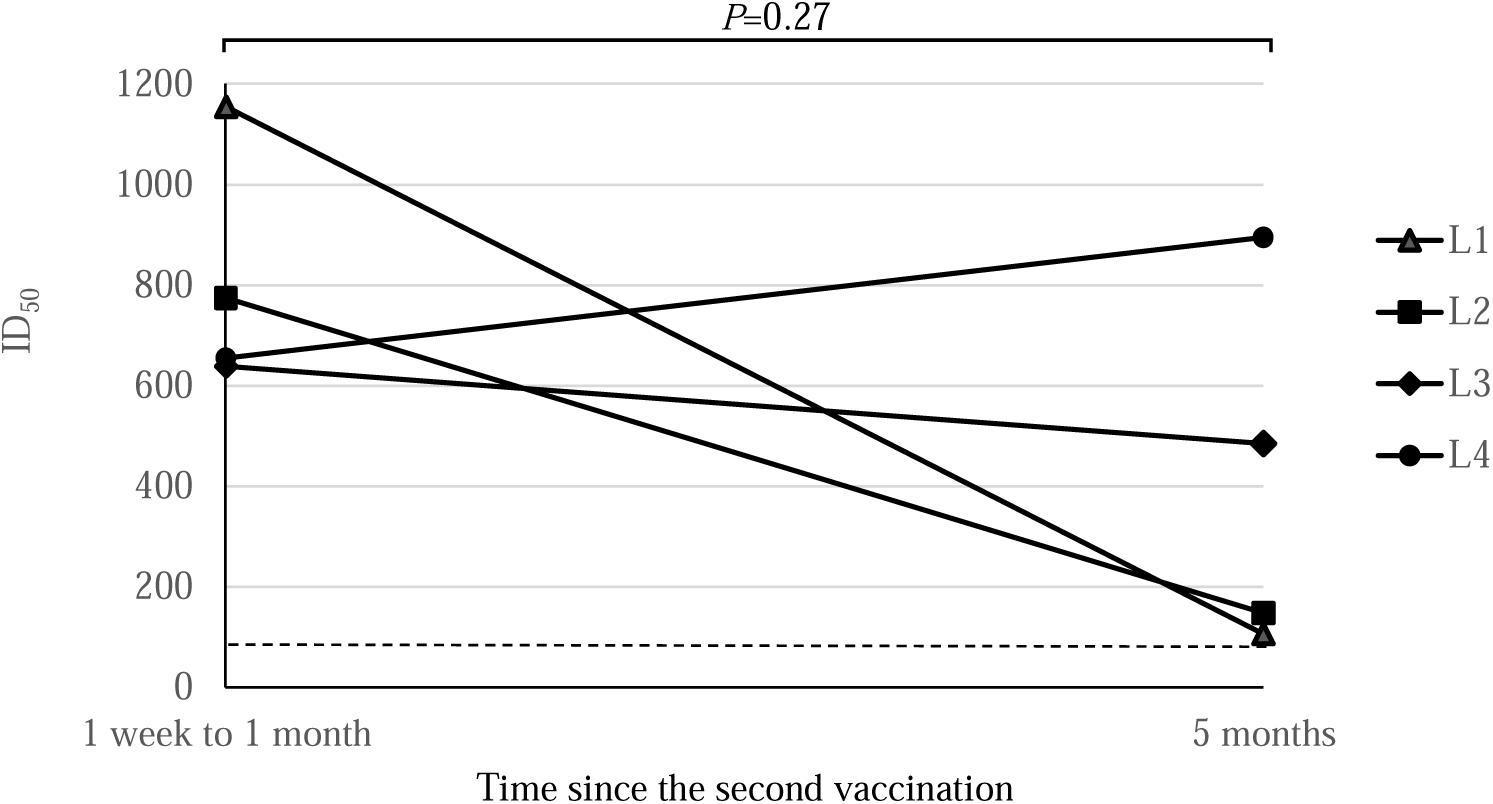
SARS-CoV-2 neutralizing antibody activity responses after the second dose of BNT162b2 vaccine in suboptimal responders.

Data are presented as 50% inhibitory dilutions (ID_50_). The dashed line indicates the cutoff for ID_50_ (17).

## Discussion

The BNT162b2 vaccine has been highly effective in preventing clinically significant COVID-19 (8-10). However, limited information is available about the effectiveness of the BNT162b2 vaccine in suboptimal responders to conventional vaccines. In this study, suboptimal responders produced adequate SARS-CoV-2 anti-spike antibodies and neutralizing antibodies.

Muik et al. (19) measured the neutralization efficacy of sera from 40 healthy German individuals immunized with the BNT162b2 vaccine and a 50% inhibitory dilution factor using pseudovirus according to the same method as was used in this study. The SARS-CoV-2 neutralizing activity of the suboptimal responders in this study 1 week to 1 month after receiving the second dose of BNT162b2 vaccine was comparable to that of the participants in the German study. All four suboptimal responders developed protective levels of neutralizing antibodies. The SARS-CoV-2 neutralizing antibody activity value in the fifth month after the second dose of BNT162b2 vaccine did not show a significant decrease in activity compared with that 1 week to 1 month after receiving the second dose of BNT162b2 vaccine. The SARS-CoV-2 anti-spike antibody titer was lower at 5 months after the second dose of BNT162b2 vaccine than at 1 week after the second dose, but the decrease was not statistically significant in suboptimal responders. These results suggest that mRNA vaccines can invoke a protective immune response even in suboptimal responders who do not develop adequate antibody titers against viruses in response to conventional vaccines against viral diseases such as mumps and measles.

Measles, rubella, chickenpox, and mumps vaccines are live attenuated vaccines that weaken the toxicity of pathogenic microorganisms, and hepatitis B vaccines are inactivated vaccines that cause pathogenic microorganisms to lose their ability to infect (20). In previous studies, approximately 2–10% of healthy individuals do not produce protective antibody levels in response to viral vaccines (21). The risk factors for low response to these conventional viral vaccines include male sex, age of >40 years, smoking habit, obesity, chronic diseases, and genetic factors. However, the factors associated with a low antibody response to SARS-CoV-2 mRNA vaccines are not well understood. Smokers, patients with cancer and hematological malignancies, and organ transplant recipients have been reported to have a lower SARS-CoV-2 anti-spike antibody response than individuals without these characteristics after receiving SARS-CoV-2 vaccines (22-26). It is intriguing that the suboptimal responders in this study who did not produce adequate antibodies against measles or mumps produced adequate SARS-CoV-2 anti-spike antibodies after receiving the BNT162b2 vaccine. They were healthy and did not have risk factors such as an organ transplant and immunosuppression. One possible reason for this discrepancy is the different mechanism of antibody production between conventional vaccines and SARS-CoV-2 mRNA vaccines.

The BNT162b2 vaccine is an mRNA vaccine that incorporates mRNA encoding spike protein (antigen) on the surface of the SARS-CoV-2 into human cells. The produced antigen directly functions as a template to induce the acquired immune system (27). On the other hand, with live attenuated vaccines or inactivated vaccines, it is necessary for recipients to process the vaccine and recognize it as an antigen. The difference in the mechanism of action of vaccines could be a factor contributing to the discrepancy in response to the BNT162b2 vaccine from the response to conventional live attenuated vaccines among suboptimal responders. Such a hypothesis should be tested in future studies. It is also unclear why suboptimal responders had higher antibody titers than normal responders in the first week after the second dose of vaccine.

Of note, the SARS-CoV-2 anti-spike antibody level is known to peak and then decrease within months after the second vaccination, which is a much shorter period than that of antibodies against live attenuated vaccines (28,29). In order to achieve adequate COVID-19 disease control, a third dose of SARS-CoV-2 mRNA vaccine is needed, which is already being administered in many countries. In view of the results of the present study, a third dose of the BNT162b2 vaccine is recommended for both suboptimal and normal responders to conventional vaccines. (28)

This study has some limitations. The number of participants was limited, and all participants were HCWs vaccinated at a single hospital. In addition, there were no suboptimal responders to inactivated vaccines (such as hepatitis B) included in this study. Multicenter studies with a larger sample size are needed to confirm the results of the present study. In addition, further immunological studies are necessary to clarify the mechanism by which humoral immunity is acquired after mRNA vaccination in suboptimal responders to conventional vaccines.

In conclusion, after two doses of the BNT162b2 vaccine, suboptimal responders to conventional antiviral vaccines developed protective levels of SARS-CoV-2 anti-spike and SARS-CoV-2 neutralizing antibodies. These results suggest that vaccination with mRNA vaccine against SARS-CoV-2 should also be recommended to suboptimal responders against conventional vaccines.

## Data Availability

All data produced in the present work are contained in the manuscript

## Ethics consideration

We carried out a prospective cohort study of 50 HCWs. This study was conducted with the approval of the Ethics Committee of Kyorin University School of Medicine (Number R02-041), and was in accordance with the Declaration of Helsinki. Informed consent was obtained from all study participants.

## Acknowledgments

We thank the HCWs of the Department of Clinical Laboratory, Kyorin University Hospital involved in the SARS-CoV-2 antibody monitoring for their valuable contribution.

